# Device-measured stationary behaviour and cardiovascular and orthostatic circulatory disease incidence: a population cohort study of 83,013 adults

**DOI:** 10.1101/2024.01.17.24301458

**Authors:** Matthew N. Ahmadi, Pieter Coenen, Leon Straker, Emmanuel Stamatakis

## Abstract

**Importance:** Previous studies have indicated that standing maybe beneficially associated with surrogate metabolic markers, while more time spent sitting has an adverse association. Studies assessing the dose-response associations of standing, sitting, and composite stationary behaviour time with cardiovascular disease (CVD) and orthostatic circulatory disease are scarce and show an unclear picture.

**Objective:** To examine associations of daily sitting, standing, and stationary time with CVD and orthostatic circulatory disease incidence

**Methods:** We used accelerometer data from 83,013 adults (mean age±SD= 61.3±7.8; Female=55.6%) from the UK Biobank to assess daily time spent sitting and standing. Major CVD was defined as coronary heart disease, heart failure, and stroke. Orthostatic circulatory disease was defined as orthostatic hypotension, varicose vein, chronic venous insufficiency, and venous ulcers.

**Results:** During 6.9 (±0.9) years of follow-up 6,829 CVD and 2,042 orthostatic circulatory disease events occurred. When stationary time exceeded 12 hrs/day orthostatic circulatory disease risk was higher by an average HR [95% CI] of 0.22 [0.16, 0.29] per hour. Every additional hour above 10 hrs/day of sitting was associated with a 0.26 [0.18, 0.36] higher risk. Standing more than 2 hrs/day was associated with an 0.11 [0.05, 0.18] higher risk for every additional 30 min/day. For major CVD, when stationary time exceeded 12 hrs/day, risk was higher by an average of 0.13 [0.10, 0.16] per hour. Sitting time was associated with a 0.15 [0.11, 0.19] higher risk per extra hour. Time spent standing was not associated with major CVD risk.

**Conclusions:** Time spent standing was not associated with CVD risk but was associated with higher orthostatic circulatory disease risk. Time spent sitting above 10 hours/day was associated with both higher orthostatic circulatory disease and major CVD risk. The deleterious associations of overall stationary time were primarily driven by sitting. Collectively, our findings indicate increasing standing time as a prescription may not lower major CVD risk and may lead to higher orthostatic circulatory disease risk.

## Introduction

Sitting and standing postures are termed collectively “stationary behaviour”, i.e. no ambulatory movement and low energy expenditure.^1^ Both postures have attracted interest as independent risk factors for cardiovascular disease (CVD) and premature mortality^2–6^. Intervention and cross-sectional studies have suggested decreasing sitting (the main component of sedentary time) and standing may improve surrogate cardiovascular outcomes such as metabolic markers (e.g. density lipoproteins, total cholesterol, triglycerides)^7,8^. However, studies assessing clinical endpoints such as CVD hospitalisation and mortality risk are very scarce and show an unclear picture for the dose-response of both sitting and standing^9,10^.

The majority of prospective sitting and standing time studies have relied on self-report measures, known for their inherent biases, eg. social desirability and recall, leading to imprecise evidence on links with cardiovascular disease incidence ^11–13^. Importantly, prior studies have not differentiated orthostatic circulatory diseases from other CVD types when assessing sitting and standing time. Postural influences on autonomic neuropathy, and strain on the vascular (e.g., haemodynamic) and musculoskeletal systems may create distinctly separate mechanistic pathways between different postures (sitting and standing) and thus orthostatic circulatory diseases versus other CVD types such as coronary heart disease and stroke^14–18^. Collectively, these limitations may have contributed to the inconclusive evidence regarding the associations of standing time with CVD risk.

Prior studies with mortality and CVD outcomes^19–21^ have also unintentionally examined stationary behaviour by using waist attached wearable devices that only measured ambulatory activities and cannot differentiate between sitting and standing. Such misclassification in these studies, which were originally aimed at examining sedentary behaviour, may have distorted dose-response estimates of sitting time since standing often occupies approximately 20% to 30% of adults’ waking times (3-5 hours per day^22,23^). Despite all these limitations of the current literature and the absence of consistent evidence with clinical endpoints^4,9^, standing has been recommended as health enhancing by clinicians and public health researchers^7,24–26^ .

In a large population sample of adults with device-based measures of posture and physical activity, we examined the prospective associations of stationary behaviour and its constituent components (sitting and standing) with major CVD incidence (coronary heart disease, stroke, and heart failure) and orthostatic circulatory disease.

## Methods

### Study participants

Participants were included from the UK Biobank Study, a prospective cohort of 502,629 participants between 40-69 years. All participants were enrolled between 2006-2010 and provided informed written consent. Ethical approval was provided by the UK’s National Health Service, National Research Ethics Service (Ref 11/NW/0382). Participants completed physical examinations by trained staff and touchscreen questionnaires.^13^

### Orthostatic circulatory disease and CVD incidence ascertainment

Participants were followed up through October 31^st^, 2021, with deaths obtained through linkage with the National Health Service (NHS) Digital of England and Wales or the NHS Central Register and National Records of Scotland (September 30^th^ to October 31^st^, 2021). Inpatient hospitalisation data were provided by either the Hospital Episode Statistics for England, the Patient Episode Database for Wales, or the Scottish Morbidity Record for Scotland (September 30^th^ 2021 for England, July 31^st^ for Scotland, and February 28^th^ 2018 for Wales). Primary care data were linked up to March 31^st^ to August 31^st^ 2017. We defined orthostatic circulatory disease events as orthostatic hypotension, varicose vein, chronic venous insufficiency, and venous ulcers ^27,28^. Major CVD was defined as coronary heart disease, stroke, and heart failure. Full methods for the assessment of orthostatic circulatory disease and CVD events and ICD-10 codes are provided in **Supplemental Table 1**.

### Sitting, standing, and non-stationary physical activity assessment

Between 2013 and 2015, 103,684 participants wore an Axivity AX3 accelerometer (Axivity Ltd, Newcastle upon Tyne, UK) on their dominant wrist for 24-hrs/day for 7 days to measure physical activity.^24^ Devices were calibrated and non-wear periods were identified according to standard procedures^29–31^. Primary exposures were daily time spent stationary (i.e. sitting and standing combined), sitting, and standing and were all classified with an accelerometer-based activity machine learning scheme that has been previously validated under free living conditions^32,33^. Briefly, this activity scheme uses features in the raw acceleration signal to identify and quantify time spent in sitting, standing, standing with movement, and walking/running in 60-second windows. Under free-living conditions, the activity classifier had a balanced accuracy (combination of sensitivity and specificity) of 88% for sitting time and 80% for standing time)^33^. We provide additional independent validation results in **Supplemental Text 1** suggesting overall balanced accuracy of 84%.

### Covariates

In line with previous analogous studies^9,34^ and known correlates of posture^23^, covariates in our analyses included age, sex, body mass index^35^, smoking status, alcohol consumption, fruit and vegetable consumption, education, self-reported parental history of CVD, prevalent major CVD or orthostatic circulatory disease events as appropriate, and cholesterol, anti-hypertensive, or diabetes medication use. All analyses were also adjusted for accelerometer measured time spent stepping. We also included mutual adjustment for sitting time and standing time in the corresponding models. Complete covariate definitions are provided in **Supplemental Table 2.**

### Analyses

We excluded participants with prevalent orthostatic circulatory disease and major CVD as appropriate, ascertained through self-report, hospital admission, and primary care linkage, as well as participants who were underweight (body mass index < 18.5 kg/m^2^), had missing covariate data, or had an event within the first 12 months following the accelerometry measurements (**Supplemental Figure 1)**.

We calculated the adjusted dose-response absolute risk using Poisson regression, age- and sex-adjusted incidence rate ratios, and crude risk percent. We used Cox proportional hazards regression models to estimate the dose-response hazard ratios (HR) with 95% CIs for orthostatic circulatory disease and CVD events, with natural splines and knots placed at the 10^th^, 50^th^, and 90^th^ percentiles for stationary, standing and sitting time distributions. Fine-Gray subdistribution method was used with non-orthostatic circulatory disease or non-CVD mortality events treated as a competing risk, where appropriate. The proportional hazards assumption was assessed using Schoenfeld Residuals, and no violations were observed. Due to an absence of prior studies assessing clinical endpoints, we used the adjusted absolute risk curves to determine reference values for the three exposures, using the data point where orthostatic circulatory disease and CVD risk became pronounced (**Figures 1-2**). The reference points were 12 hours/day, 10 hours/day, and 2.5 hours/day for stationary, sitting, and standing time, respectively. Departure from linearity was assessed by a Wald test examining the null hypothesis that the coefficient of the second spline was equal to zero. We calculated E-values to estimate the plausibility of bias from unmeasured confounding. The E-values indicate the required magnitude of the association unmeasured confounders to reduce findings to null. To provide conservative E-value point estimates, we assessed the minimal dose, defined as the duration of each exposure associated with 50% of the highest HR (‘minimum harmful dose’).

**Figure 1:**
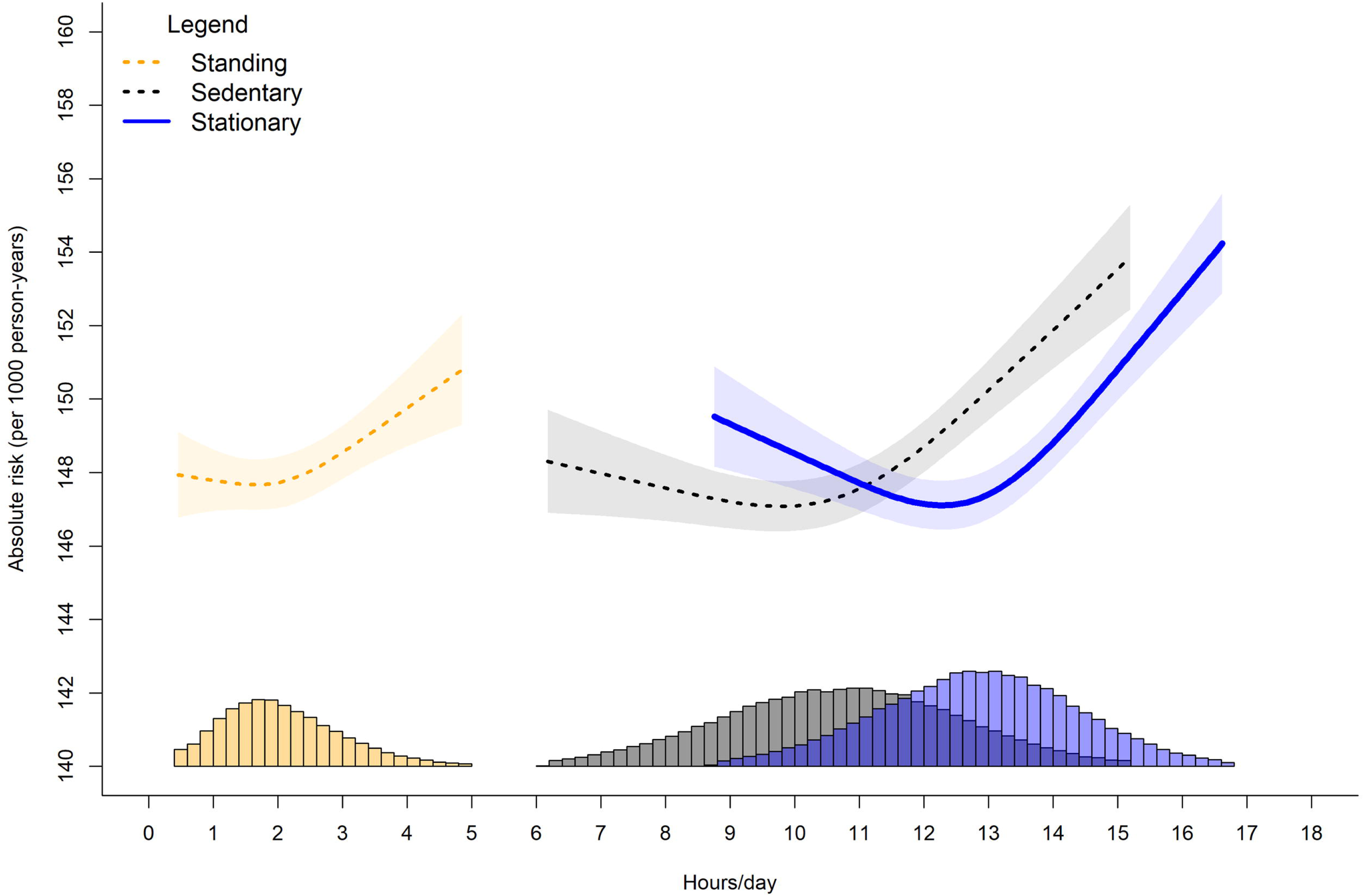
Adjusted absolute risk of stationary, standing, and sitting time with orthostatic circulatory disease incidence. Legend: Adjusted for age, sex, ethnicity, smoking history, alcohol consumption, time spent walking, mutual adjustment for time spent standing and sitting, education, diet, family history of CVD, prevalent CVD incidence, and medication use.

**Figure 2:**
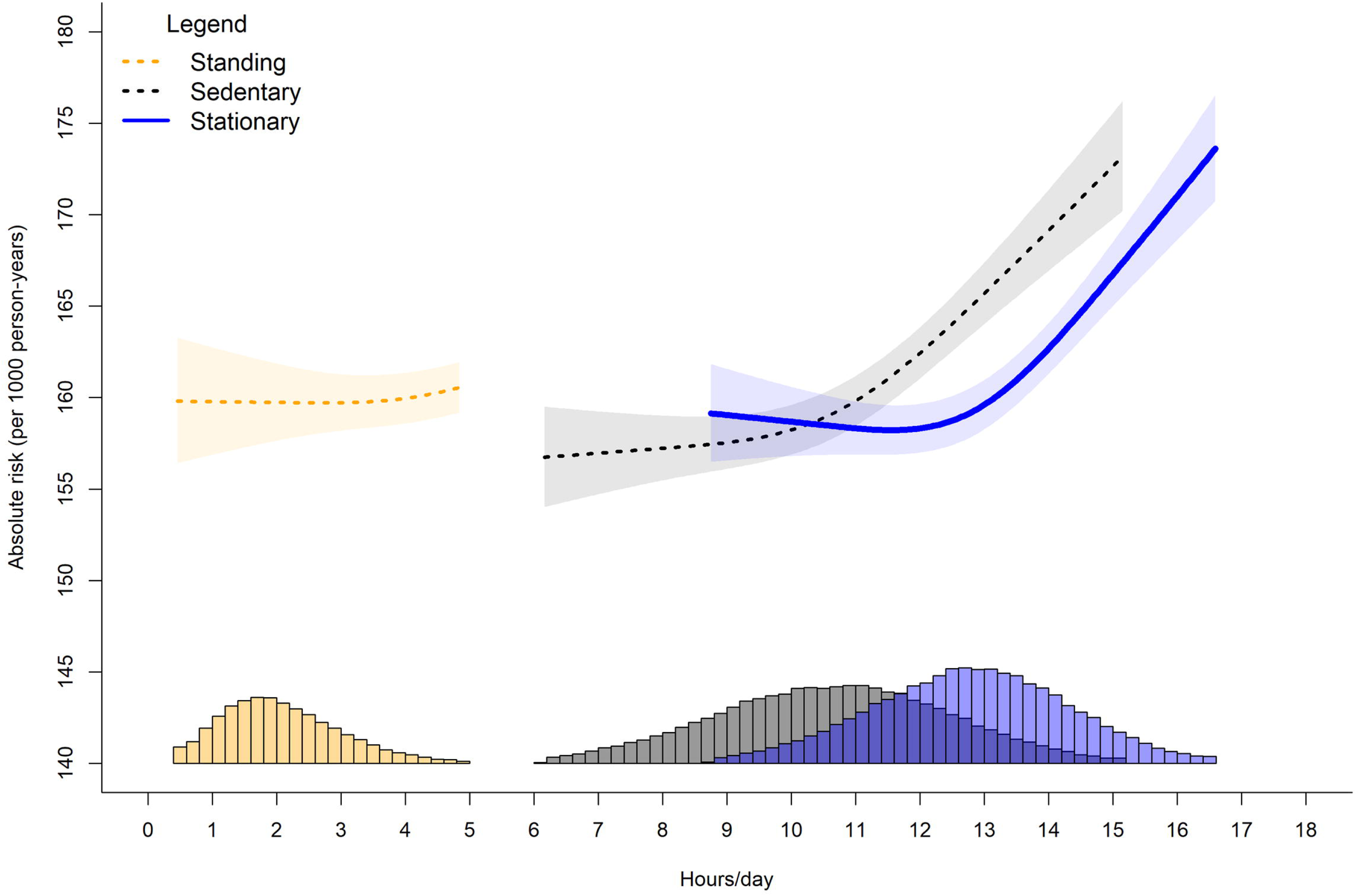
Adjusted absolute risk of stationary, standing, and sitting time with major cardiovascular disease incidence. Legend: Adjusted for age, sex, ethnicity, smoking history, alcohol consumption, time spent walking, mutual adjustment for time spent standing and sitting, education, diet, family history of CVD, prevalent orthostatic incidence, and medication use.

To assess the influence of residual confounding, we used a negative control outcome of deaths and hospitalisation from accidents (excluding accidents that may be associated with physical activity. i.e. cycling, and falls incidence, or self-harm), an outcome that does not have a mechanistic link to stationary behaviour. Negative controls can improve causal inference by illustrating pervasive bias and confounding. If the negative control has a similar association pattern as the primary outcomes, then it is more plausible associations are due to bias and confounding than causal mechanisms^36^. We conducted sensitivity analyses to minimise bias attributable to reverse causation by: 1) exclusion of participants who were obese (body mass index >30 kg/m^2^); 2) exclusion of participants reporting fair or poor health; or 3) those with an event within the first 24 months of follow-up.

We performed all analyses using R statistical software. We reported this study as per the Strengthening the Reporting of Observational Studies in Epidemiology (STROBE) guideline (see **STROBE Statement** in the Supplement).

## Results

Our analytic sample for orthostatic circulatory disease incidence included 83,013 participants (average age [sd]= 61.3 (7.8) years; 55.6% female) followed-up for an average of 6.9 ± 0.9 years with 2,042 events. Our CVD incidence sample included 75,897 participants with 6,829 events. Mean (SD) time spent stationary, standing and sitting was 12.8 (1.6) hours/day, 2.1 (0.9) hours/day, and 10.7 (1.9) hours/day, respectively. Participants spent an average of 71.3 minutes/day ambulatory (walking/running) at any intensity and 64.0% of the participants were not smokers. Participant characteristics by stationary behaviour daily duration is provided in **Table 1**. Participant characteristics by standing and sitting daily duration are presented in **Supplemental Tables 3** and **4**. Participants in the higher quartiles of sitting were more likely to be male and have tertiary education, whilst participants in higher quartiles of standing were more likely to be female and less educated.

**Table 1:**
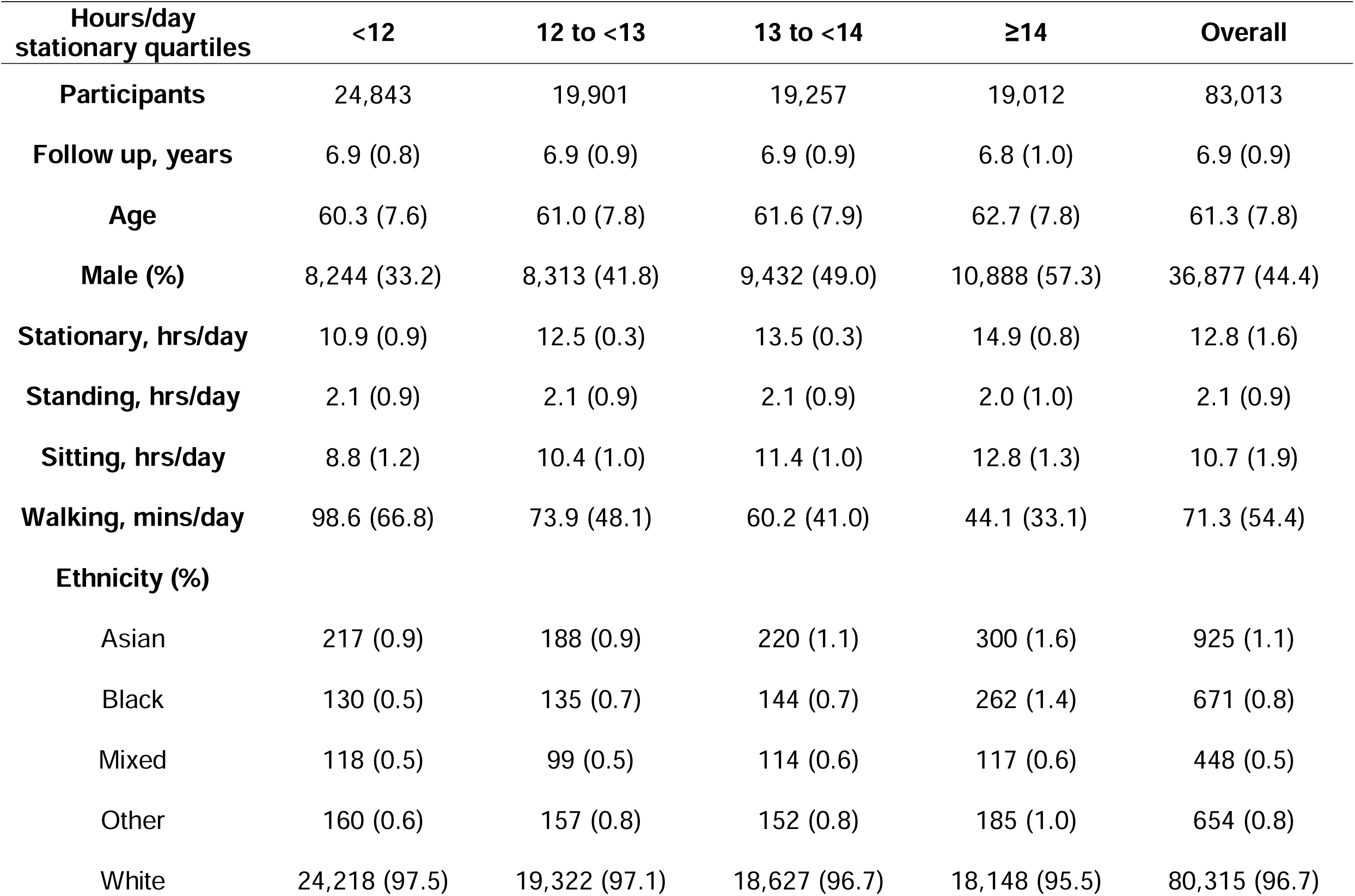
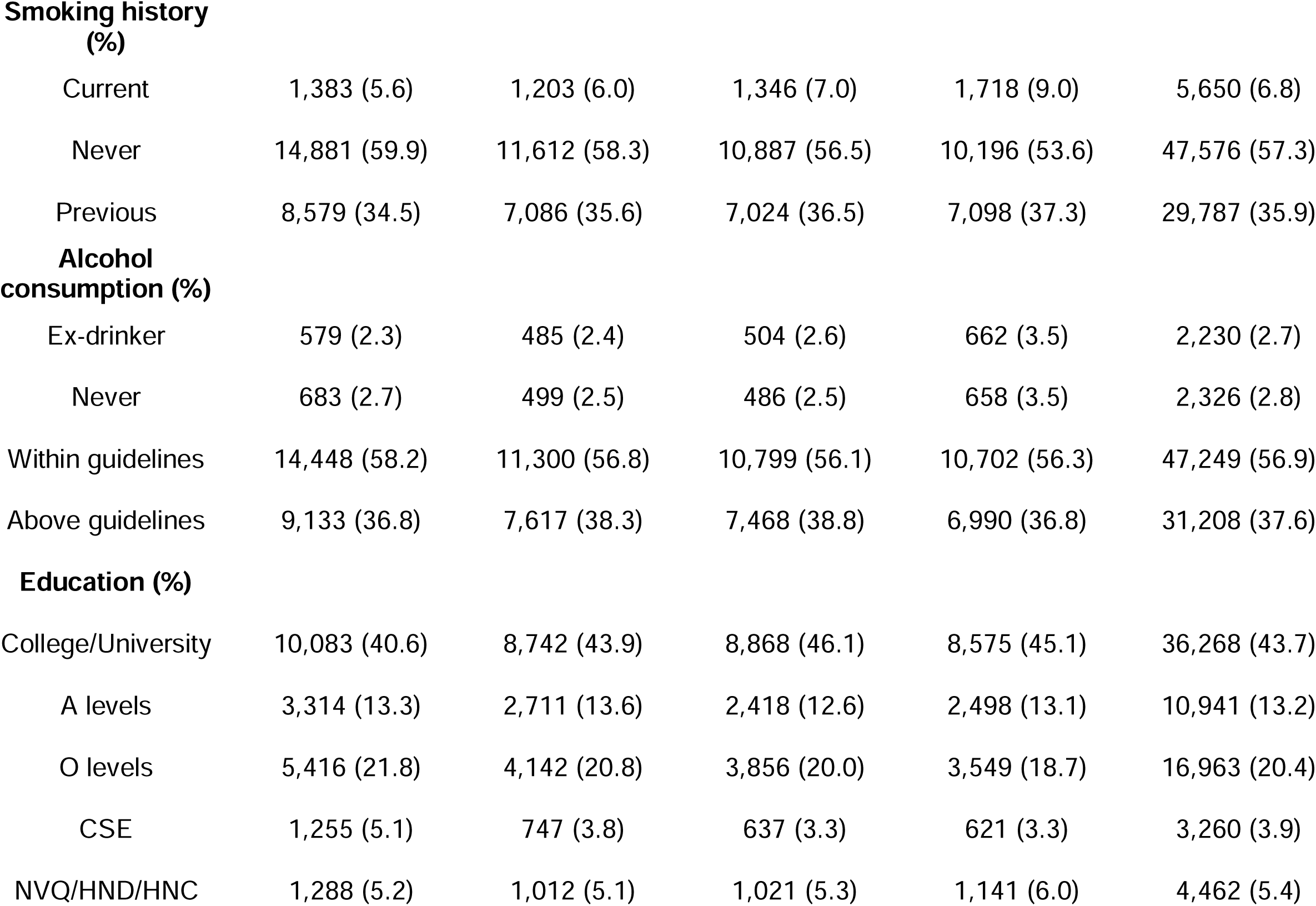
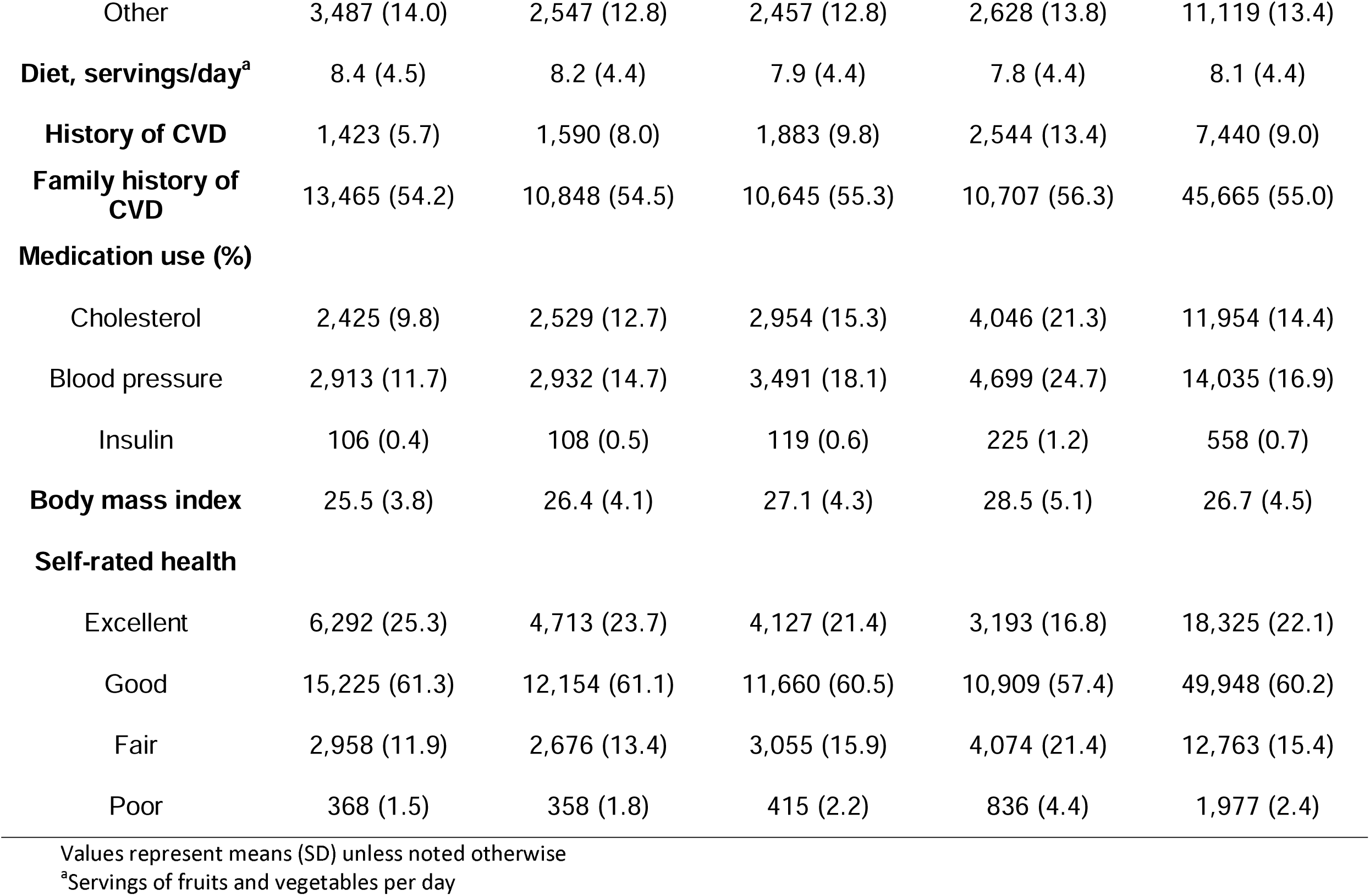
Participant descriptive characteristics by stationary time.

| **Hours/day**<br>**stationary quartiles** | **<12** | **12 to <13** | **13 to <14** | **≥14** | **Overall** |
| --- | --- | --- | --- | --- | --- |
| **Participants** | 24,843 | 19,901 | 19,257 | 19,012 | 83,013 |
| **Follow up, years** | 6.9 (0.8) | 6.9 (0.9) | 6.9 (0.9) | 6.8 (1.0) | 6.9 (0.9) |
| **Age** | 60.3 (7.6) | 61.0 (7.8) | 61.6 (7.9) | 62.7 (7.8) | 61.3 (7.8) |
| **Male (%)** | 8,244 (33.2) | 8,313 (41.8) | 9,432 (49.0) | 10,888 (57.3) | 36,877 (44.4) |
| **Stationary, hrs/day** | 10.9 (0.9) | 12.5 (0.3) | 13.5 (0.3) | 14.9 (0.8) | 12.8 (1.6) |
| **Standing, hrs/day** | 2.1 (0.9) | 2.1 (0.9) | 2.1 (0.9) | 2.0 (1.0) | 2.1 (0.9) |
| **Sitting, hrs/day** | 8.8 (1.2) | 10.4 (1.0) | 11.4 (1.0) | 12.8 (1.3) | 10.7 (1.9) |
| **Walking, mins/day** | 98.6 (66.8) | 73.9 (48.1) | 60.2 (41.0) | 44.1 (33.1) | 71.3 (54.4) |
| **Ethnicity (%)** |  |  |  |  |  |
| Asian | 217 (0.9) | 188 (0.9) | 220 (1.1) | 300 (1.6) | 925 (1.1) |
| Black | 130 (0.5) | 135 (0.7) | 144 (0.7) | 262 (1.4) | 671 (0.8) |
| Mixed | 118 (0.5) | 99 (0.5) | 114 (0.6) | 117 (0.6) | 448 (0.5) |
| Other | 160 (0.6) | 157 (0.8) | 152 (0.8) | 185 (1.0) | 654 (0.8) |
| White | 24,218 (97.5) | 19,322 (97.1) | 18,627 (96.7) | 18,148 (95.5) | 80,315 (96.7) |
| **Smoking history (%)** |  |  |  |  |  |
| Current | 1,383 (5.6) | 1,203 (6.0) | 1,346 (7.0) | 1,718 (9.0) | 5,650 (6.8) |
| Never | 14,881 (59.9) | 11,612 (58.3) | 10,887 (56.5) | 10,196 (53.6) | 47,576 (57.3) |
| Previous | 8,579 (34.5) | 7,086 (35.6) | 7,024 (36.5) | 7,098 (37.3) | 29,787 (35.9) |
| **Alcohol consumption (%)** |  |  |  |  |  |
| Ex-drinker | 579 (2.3) | 485 (2.4) | 504 (2.6) | 662 (3.5) | 2,230 (2.7) |
| Never | 683 (2.7) | 499 (2.5) | 486 (2.5) | 658 (3.5) | 2,326 (2.8) |
| Within guidelines | 14,448 (58.2) | 11,300 (56.8) | 10,799 (56.1) | 10,702 (56.3) | 47,249 (56.9) |
| Above guidelines | 9,133 (36.8) | 7,617 (38.3) | 7,468 (38.8) | 6,990 (36.8) | 31,208 (37.6) |
| **Education (%)** |  |  |  |  |  |
| College/University | 10,083 (40.6) | 8,742 (43.9) | 8,868 (46.1) | 8,575 (45.1) | 36,268 (43.7) |
| A levels | 3,314 (13.3) | 2,711 (13.6) | 2,418 (12.6) | 2,498 (13.1) | 10,941 (13.2) |
| O levels | 5,416 (21.8) | 4,142 (20.8) | 3,856 (20.0) | 3,549 (18.7) | 16,963 (20.4) |
| CSE | 1,255 (5.1) | 747 (3.8) | 637 (3.3) | 621 (3.3) | 3,260 (3.9) |
| NVQ/HND/HNC | 1,288 (5.2) | 1,012 (5.1) | 1,021 (5.3) | 1,141 (6.0) | 4,462 (5.4) |
| Other | 3,487 (14.0) | 2,547 (12.8) | 2,457 (12.8) | 2,628 (13.8) | 11,119 (13.4) |
| **Diet, servings/day<sup>a</sup>** | 8.4 (4.5) | 8.2 (4.4) | 7.9 (4.4) | 7.8 (4.4) | 8.1 (4.4) |
| **History of CVD** | 1,423 (5.7) | 1,590 (8.0) | 1,883 (9.8) | 2,544 (13.4) | 7,440 (9.0) |
| **Family history of CVD** | 13,465 (54.2) | 10,848 (54.5) | 10,645 (55.3) | 10,707 (56.3) | 45,665 (55.0) |
| **Medication use (%)** |  |  |  |  |  |
| Cholesterol | 2,425 (9.8) | 2,529 (12.7) | 2,954 (15.3) | 4,046 (21.3) | 11,954 (14.4) |
| Blood pressure | 2,913 (11.7) | 2,932 (14.7) | 3,491 (18.1) | 4,699 (24.7) | 14,035 (16.9) |
| Insulin | 106 (0.4) | 108 (0.5) | 119 (0.6) | 225 (1.2) | 558 (0.7) |
| **Body mass index** | 25.5 (3.8) | 26.4 (4.1) | 27.1 (4.3) | 28.5 (5.1) | 26.7 (4.5) |
| **Self-rated health** |  |  |  |  |  |
| Excellent | 6,292 (25.3) | 4,713 (23.7) | 4,127 (21.4) | 3,193 (16.8) | 18,325 (22.1) |
| Good | 15,225 (61.3) | 12,154 (61.1) | 11,660 (60.5) | 10,909 (57.4) | 49,948 (60.2) |
| Fair | 2,958 (11.9) | 2,676 (13.4) | 3,055 (15.9) | 4,074 (21.4) | 12,763 (15.4) |
| Poor | 368 (1.5) | 358 (1.8) | 415 (2.2) | 836 (4.4) | 1,977 (2.4) |

Values represent means (SD) unless noted otherwise
<sup>a</sup>Servings of fruits and vegetables per day

## Absolute risk

**Supplemental Tables 5-6** present the crude risk and partially adjusted incidence rate ratios for orthostatic circulatory disease and major CVD events by quartiles of each exposure. For orthostatic circulatory disease, participants who had <12 hours/day of stationary time had a crude risk of 2.49% (95% CI= 2.46%, 2.52%), whereas ≥14 hours/day had a crude risk of 5.22% (5.14%, 5.29%). Corresponding values for CVD are 2.70% (2.67%, 2.74%) for <12 hours/day and 5.13% (5.05%, 5.20%) for ≥14 hours/day. The adjusted absolute risk dose-responses per 1,000 person years are shown in **Figures 1-2**. For CVD and orthostatic circulatory disease incidence, the dose-response was non-linear for stationary and sitting time with risk becoming more pronounced after approximately 12 hours/day and 10.5 hours/day, respectively. For standing time, the risk became more pronounced after 2 hours/day for orthostatic circulatory disease, however we observed no changes in CVD incidence risk between 1 hour/day (e.g. 160 [95% CI= 157, 163] events/1,000 person-years) through 4 hours/day (160 [158, 162] events/1,000 person-years). For CVD incidence, more standing time was not associated with higher absolute risk.

### Multivariable adjusted associations for orthostatic circulatory disease

When stationary time exceeded the 12 hours/day referent data point, risk increased by an average HR of 0.22 (95% CI= 0.16, 0.29) with every 1 hour increment (**Figure 3**). Similarly, every 1 hour increment of sitting time above the reference data point of 10 hours/day (referent) was associated with an average HR increase of 0.26 (0.18, 0.36). For standing time, compared to the referent 2 hours/day, every 30 minute increment above 2 hours/day was associated with an average HR increase of 0.11 (0.05, 0.18). For all three exposures, we observed a null to weak protective association for time spent below the referent value. For example, 9 hours/day sitting and 1.5 hours/day standing was associated with an HR of 0.96 (0.93, 0.99) and 0.94 (0.90, 0.99).

**Figure 3:**
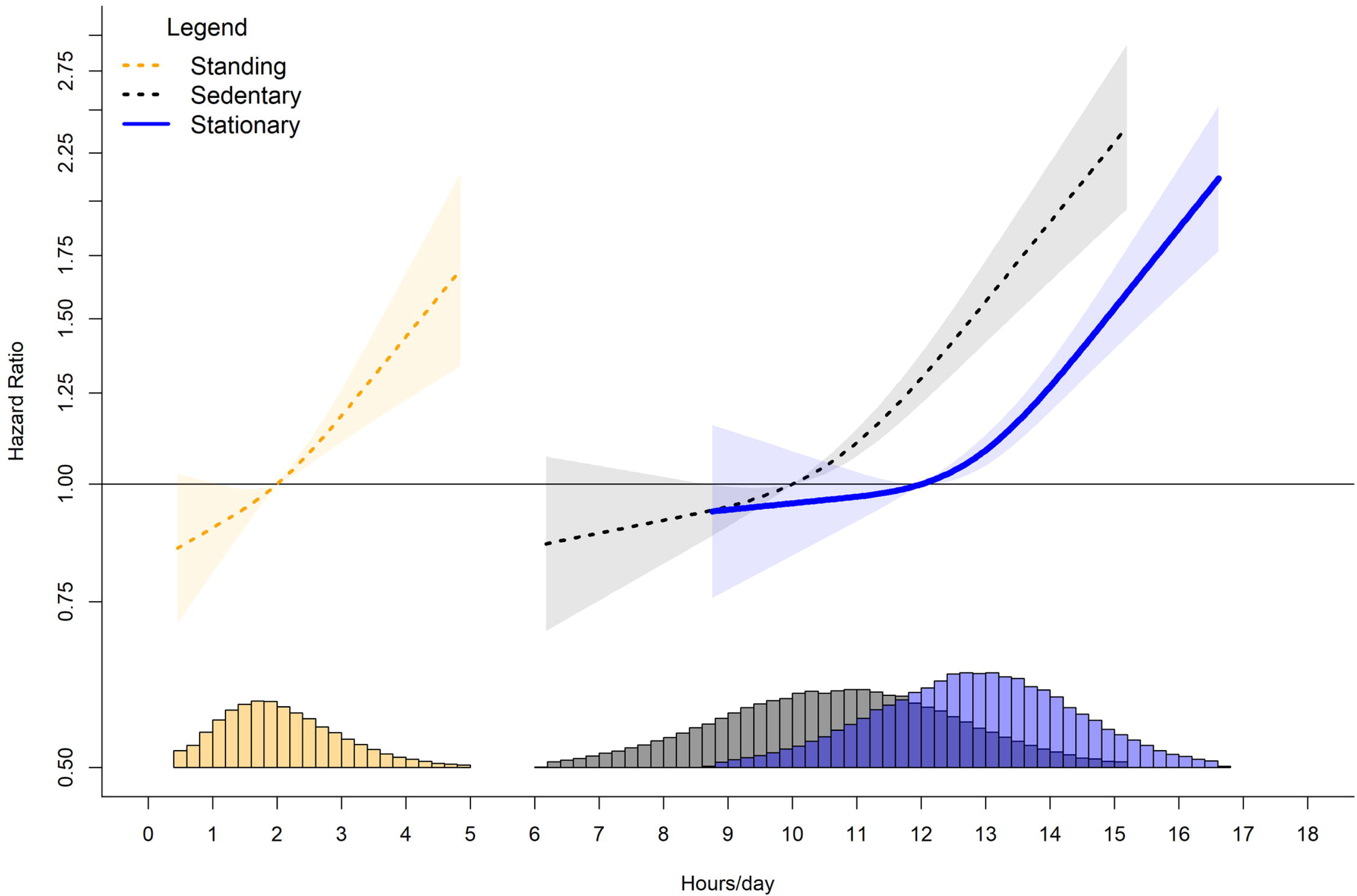
Dose-response associations of stationary, standing, and sitting time with orthostatic circulatory disease incidence. Legend: Adjusted for age, sex, ethnicity, smoking history, alcohol consumption, time spent walking, mutual adjustment for time spent standing and sitting, education, diet, family history of CVD, prevalent CVD incidence, and medication use.

### Multivariable adjusted associations for major cardiovascular disease incidence

When stationary time exceeded 12 hours/day, we observed higher CVD risk in a linear fashion (**Figure 4**). Every 1 hour increment in stationary time above 12 hours/day was associated with an average HR increase of 0.13 (0.10, 0.16). We observed lower CVD risk when stationary time was below 12 hours/day. For example, at 9 hours/day we observed an HR of 0.87 (0.78, 0.96). We also observed a linear association for sitting time when the daily duration was above 10 hours/day. Every 1 hour increment above 10 hours/day associated with an average HR increase of 0.15 (0.11, 0.19). When sitting time was below 10 hours/day we observed lower CVD risk (e.g. 9 hours/day of sitting time had an HR of 0.95 (0.93, 0.97)). Contrary to sitting time, more time spent standing was not associated with a higher CVD risk. Overall, there was no association for higher or lower CVD risk throughout the range of standing duration.

**Figure 4:**
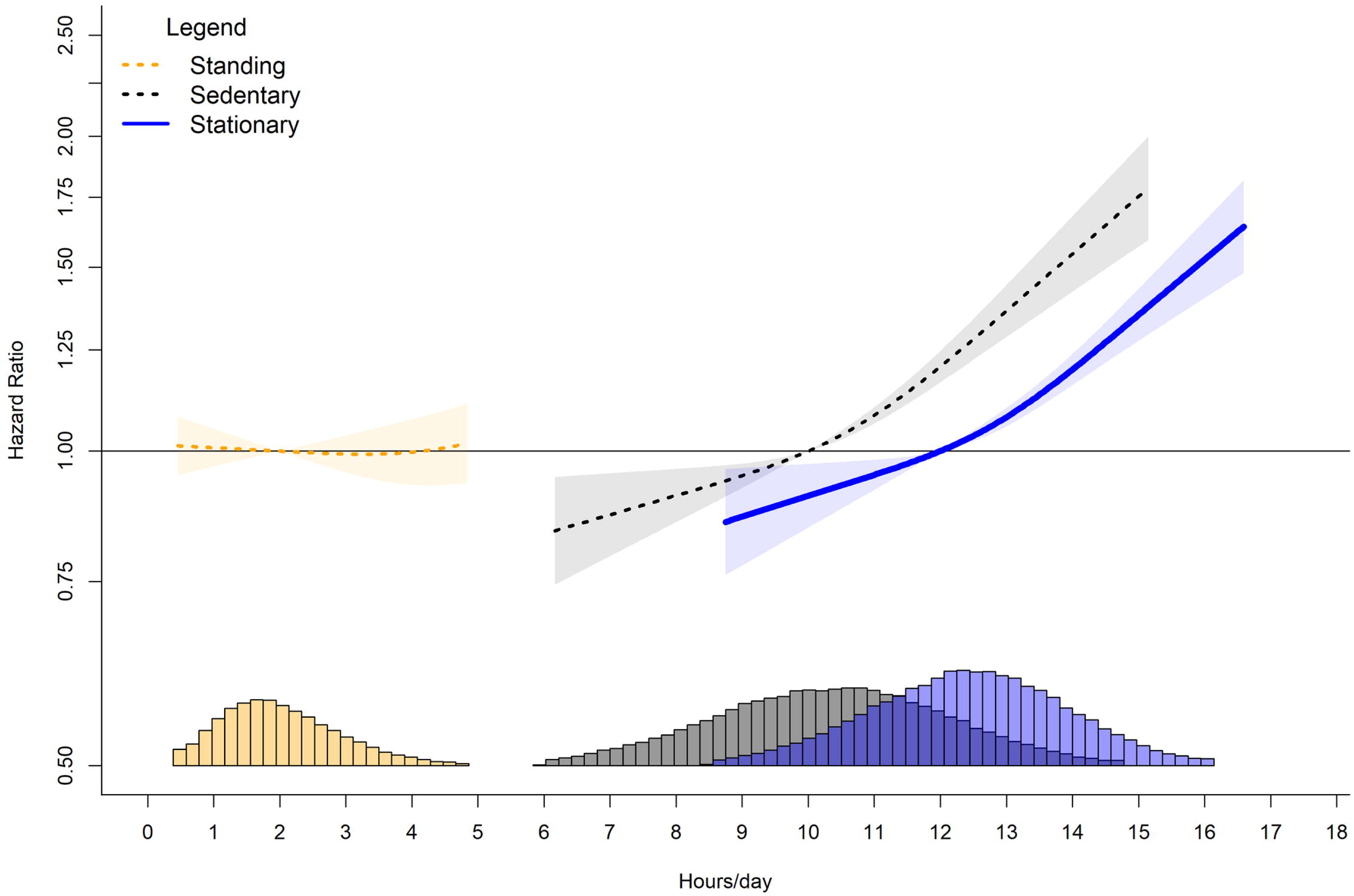
Dose-response association of stationary, standing, and sitting time with major cardiovascular disease incidence. Legend: Adjusted for age, sex, ethnicity, smoking history, alcohol consumption, time spent walking, mutual adjustment for time spent standing and sitting, education, diet, family history of CVD, prevalent orthostatic incidence, and medication use.

### Additional and sensitivity analyses

The association pattern for orthostatic circulatory disease and CVD incidence remained consistent after exclusion of participants who were: 1) obese; 2) reported fair or poor health; 3) or had an event within the first 24 months of follow-up (**Supplemental Figures 2-3**). Negative control outcomes and E-values analyses for the minimum harmful dose indicated that residual and unmeasured confounding had minimal impact on the findings. Specifically, with the negative control outcome, associations for standing were non-significant with wide 95% CIs, and sitting behaviour point estimates were inconsistent with the main analysis showing a U-shaped association (**Supplemental Figure 4**). The E-values suggest a substantial degree of unmeasured confounding would be required to reduce our observed associations at the minimum harmful dose for orthostatic circulatory disease (e.g., an association of 2.01 to 2.79) and CVD incidence (1.97 to 2.13) to null (**Supplemental Table 7**).

## Discussion

In a largest wearables-based study of stationary time and its constituent components of standing and sitting time in > 83,000 adults we observed a linear association for higher orthostatic circulatory disease risk from increased standing time with no protective association for CVD risk. After approximately 10.5 hours/day, we observed a deleterious association of increased sitting time with higher risk of both orthostatic circulatory disease and CVD risk. This calls to question current intervention strategies that focus on only replacing sitting with standing time without increasing physical activity^37^.

### Orthostatic circulatory disease risk

stationary time as well as its constituent postures sitting and standing behaviours were all associated with increased risk of orthostatic circulatory disease. For stationary time, risk increased by an average 22% with every 1 hour increment, above 12 hours/day. For sitting time, above 10 hours/day, risk increased by an average 26% with every 1 hour. For standing time, risk increased by an average of 11% with every 30 minute increment above 2 hours/day. The pattern of the dose-response relationship appears similar for sitting and standing (Fig 1) suggesting that a common aspect of sitting and standing, i.e. absence of ambulatory movement, is likely to be important in the mechanistic pathway for orthostatic circulatory disease. The lack of muscle movement during stationary time may result in a reduced venous return by skeletal muscle contraction and pumps contributing to venous pooling, causing orthostatic circulatory problems^38^. Therefore, a key implication of our finding is that non-stationary movement (e.g., walking, cycling, or other physical activities involving some degree of movement) is important to reduce orthostatic circulatory disease risk, aligning with current public health messaging to ‘move more’^39^. If confirmed to be causal in future randomised control trials, our findings would have implications for patient care among those at high risk of CVD. Public health strategies that promote standing as a sufficient substitute to overcome the cardiovascular health risks of sitting (e.g., as seen by common advice to adopt standing desks in office environments) may not achieve their goal.

Our findings also suggest the dose-risk association between stationary time and orthostatic circulatory disease is non-linear, with no association for lower risk below inflexion points of approximately 10 hours of sitting/day and 2 hours of standing/day. This suggests simple messaging to ‘sit less’ may not be optimal, as that would not lower the risk for those currently accumulating less than 10 hrs a day and may even increase risk of musculoskeletal^14^ and circulatory issues by increasing the time spent standing. The non-linear associations we detected suggest that up to a certain level, neither sitting nor standing are harmful for orthostatic conditions, suggesting that a there may be a healthy balance between these two behaviours. This balance likely varies among individuals depending on co-morbidities, overall health status, and daily physical activity levels^40,41^.

### Cardiovascular disease risk

Stationary and sitting behaviour were both associated with increased risk of CVD above certain thresholds. For stationary time, risk increased by an average of 13% with every 1 hour increment above the reference 12 hours/day. For sitting time, risk increased by an average of 15% with every 1 hour increment above 10 hours/day,. Standing time was not associated with CVD risk. The higher CVD risk we observed with sitting time is similar in magnitude to the associations reported in prior studies of sitting and CVD outcomes^2,34,42^. The pattern of dose-risk relationship appears similar for sitting and overall stationary but not for standing (Figs 2 and 4), suggesting that the sitting component is driving these associations, rather than the absence of movement. There are additional possible mechanisms that are unique to sitting. For example, the lower cumulative energy expenditure of sitting and the muscular and musculoskeletal system engagement during standing^43^ may partly explain the differential effects of the two postures. Although standing time was not associated with higher CVD risk, we did not observe a protective association. Collectively, the implications of our findings for public health messaging are supportive of current messages^39^ encouraging sitting reductions for CVD health, however they do not support increasing standing time alone as a mitigation strategy cited in some guidelines^44^.

### Strengths and limitations

A key strength of this study was that, unlike previous device-based studies, we were able to separately examine the components of stationary behaviours, enabling the estimation of risks associated with sitting versus standing, two postures that are underpin by different mechanisms and both have public health an clinical importance. We utilised the largest wearables -based data resource with rich contextual and linkage to health outcomes information. The wrist placement improves translation of our findings into public health messaging and immediate uptake by users of consumer level wrist wearables. Among the general public that tracks and provides feedback on time spent sitting, standing, and active throughout a day. Limitations of our study included the potential misclassification of posture and movement that is inherent to wrist worn devices, although our daily estimates are similar to sitting and standing time assessed from the gold standard of wearables postural assessment, thigh-worn devices, in other UK cohorts^23,45^. The observational design of our study precludes us from making causal interpretations. We cannot rule out the presence of unmeasured confounding, although our E-values indicated that unmeasured confounders would need to have to have a very strong association with the exposure and outcomes for the observed associations to be null. The UK Biobank had a low response rate, however previous work has shown that poor representativeness does not materially influence the associations between lifestyle risk factors and noncommunicable disease risk^46^.

## Conclusion

The deleterious associations of stationary time with CVD and orthostatic circulatory disease we observed were primarily a consequence of time spent sitting. More time spent standing was not associated with CVD risk but was associated with substantially higher risk of orthostatic circulatory disease. Collectively, our findings are supportive of clinical and public health strategies to curtail excessive sitting time as an important risk factor for major CVD. However, standing time alone may not be a sufficient mitigation strategy for lower CVD risk, and may lead to a higher risk of circulatory conditions.

## Data Availability

All data produced in the present study are available upon application to the UK BioBank.

https://www.ukbiobank.ac.uk/

